# Sexual Gender-Based Violence among Adolescent Girls and Young Women during COVID-19 Pandemic, Mid-Eastern Uganda

**DOI:** 10.1101/2023.09.11.23295394

**Authors:** Patience Mwine, Benon Kwesiga, Richard Migisha, Juliet Cheptoris, Daniel Kadobera, Lilian Bulage, Edirisa J. Nsubuga, Peter Mudiope, Alex R. Ario

## Abstract

**Background:** Global studies indicate that sexual gender based violence (SGBV) may increase during pandemics including the COVID-19. The Mid-Eastern region in Uganda was of a concern due to high prevalence of intimate partner sexual violence among adolescent girls and young women (AGYW) (13% in 2016). Due to limited data, we investigated factors associated with SGBV among AGYW during the COVID-19 pandemic in Eastern Uganda, April 2022.

**Methods:** We line listed all AGYW 10-24 years who obtained SGBV services at ten high-volume health facilities from March 2020 to December 2021, the main COVID-19 period in Uganda. We conducted a case-control study among these AGYW. A case was ≥1 SGBV episode experienced by an AGYW aged 10-24 years residing in Tororo and Busia Districts. For every randomly-selected case from the health facility line list, we identified two neighbourhood-matched AGYW controls who reported no SGBV. We interviewed 108 and 216 controls on socio-demographics, socio-economics, and SGBV experiences during COVID-19. We conducted logistic regression to obtain adjusted odds ratios and confidence intervals.

**Results:** Among 389 SGBV cases, the mean age was 16.4 (SD± 1.6: range 10-24) years, and 350 (90%) were 15-19 years. Among 108 cases interviewed, 79 (73%) reported forced sex. Most (73; 68%) knew the perpetrator. In multivariate analysis, self-reported SGBV before the COVID-19 period [aOR=5.8, 95%CI: 2.8-12] and having older siblings [aOR=1.9, 95%:CI 1.1-3.4] were associated with SGBV during the period. Living with a family that provided all the basic needs was protective [aOR=0.42, 95%: CI 0.23-0.78].

**Conclusion:** Previous SGBV experiences and family dynamics, such as having older siblings, increased the odds of SGBV during the COVID-19 pandemic in Uganda. Conversely, a supportive family environment was protective. Identifying, supporting, and enacting protective interventions for existing SGBV victims and socioeconomically vulnerable AGYW could reduce the burden of SGBV during similar events.

## Introduction

Sexual gender-based violence (SGBV) is defined as “any sexual act, attempt to obtain a sexual act, or other act directed against a person’s sexuality using coercion, by any person regardless of their relationship to the survivor, in any setting” (1). SGBV is a significant public health concern with multifaceted consequences for adolescent girls and young women (AGYW), including increased risks of HIV transmission, mother-to-child HIV transmission, unwanted pregnancies, school dropouts, mental health disorders and socio-economic difficulties (2).

In Uganda, approximately 22% of women aged 15-49 experience SGBV at some point in their lives (3). However, AGYW are disproportionately affected by sexual violence. The 2018 Violence against Children Survey (VAC) revealed that one in three girls aged 18-24 years experienced sexual violence during their childhood, while one in four girls age 13-17 years reported recent sexual violence (past 12 months) (4). Women reporting SGBV receive essential services, including timely Post Exposure Prophylaxis (PEP) and emergency contraception within 72 hours, as well as the necessary medical attention for any injuries or wounds sustained (5). However, the implementation of COVID-19 restrictions aimed at controlling the pandemic resulted in disruptions in access to post violence essential services (6).

From March 2020 to January 2022 in Uganda, schools were closed as part of the COVID-19 pandemic response. Anecdotal reports from Uganda Police and an analysis of national SGBV program data suggested that SGBV cases had increased during the COVID-19 pandemic, especially during the two lockdown periods in 2020 and 2021(7). Both the prolonged period spent out of school and the implementation of multiple lockdowns, leading to girls’ exposure to perpetrators within homes and neighborhoods during times they would otherwise have been elsewhere, were suggested as possible causes (8). A notable 33% increase in teenage pregnancies during 2020 and 2021 compared to 2019 indicated a potential rise in sexual gender-based violence (SGBV) during the COVID-19 pandemic period; analysis of Health Information Management System HMIS data (P.M., unpublished data). The Mid-Eastern Region, known for highest prevalence of sexual violence among AGYW 15-24 years (13.3%) according to Uganda Population-based HIV Impact Assessment (UPHIA) 2016 (9), was of particular concern . There was limited information on factors associated with SGBV among adolescent girls and young women during the COVID-19 pandemic. We evaluated SGBV services and determined the factors associated with SGBV among AGYW during the COVID-19 pandemic in Mid-Eastern Region to provide information for interventions and future prevention measures for SGBV.

## Methods

### Study design and setting

We employed a mixed-methods approach, including a descriptive, qualitative, and case-control study among AGYW aged 10-24 years in Tororo and Busia districts, Mid-Eastern Region. These two districts are in Eastern Uganda, bordering Kenya. Both have active trading activities and border points with many transit truck drivers. These districts were selected because they reported the highest number of teenage pregnancies during the COVID-19 period in Mid-Eastern Region (10).

### Case definition and finding

We selected ten high-volume health facilities providing SGBV care, including one district hospital, one Health Center IV, and three high-volume Health Center IIIs from each of the two districts. We abstracted health facility data for AGYW (aged 10-24 years) recorded in the SGBV register from March 2020 to December 2021 during 20th March to 2^nd^ April, 2022. Health care workers record information for SGBV survivors who are identified through self-reporting or from routine screening for SGBV, including screening that occurs as part of HIV care and treatment clinics and in antenatal care clinics. We obtained information on the visit date, socio-demographic characteristics, and patient management, including HIV post-exposure prevention (PEP), family planning, outcome, and follow-up. Additional information on PEP provision was obtained from the PEP registers.

### Descriptive epidemiology

We described the SGBV cases by place and person and the quality of SGBV services received by the survivors. Using district sub-county population estimates (District records-un-published data) as the denominator, we computed SGBV incidence rates per 100 individuals in each sub-county using health facility data. The numerator included recorded SGBV cases among AGYW aged 10-24 years from March 2020 to December 2021.

### Case-control study to identify factors associated with sexual gender-based violence during the COVID pandemic period, Tororo and Busia District, Uganda

The SGBV incidence rates (IR) among sub-counties ranged from 0.1% to 6%. Among the 11 sub-counties with IR ≥3%, we randomly selected six (three from each district) to serve as sites for the case-control study. These included Mukuju, Western division, and Usukuru sub-counties from Tororo district and Masafu, Lumino, and Buyanga sub-counties from Busia District (Figure 1).

**Figure 1.**
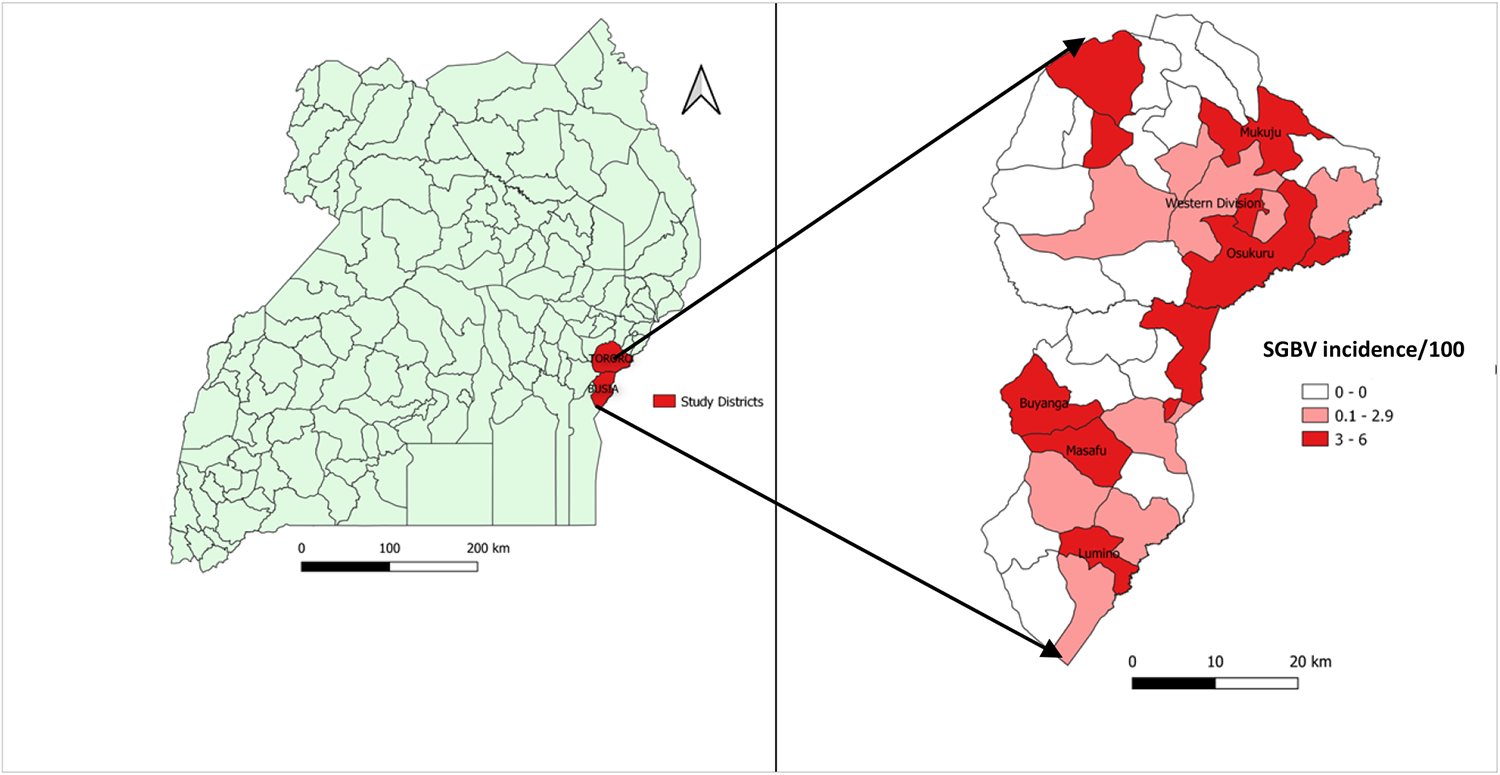
Sexual gender-based violence incidence rate among adolescent girls and young women in Busia and Tororo Districts, March 2020-December 2021.

We conducted a neighbourhood-matched case-control study to determine factors associated with SGBV among AGYW during March 2020 to December 2021, covering the highest-burden phases of the COVID-19 pandemic in Uganda. We defined a case as an AGYW aged 10-24 years who reported at least one episode of SGBV to any of 10 high-volume health facilities in Tororo and Busia Districts during this time. For every case, we identified two neighbourhood-matched AGYW controls who reported no SGBV during the same period, from 20th March to 2^nd^ April, 2022.

We assumed a self-reported reduction in family income during the pandemic was a significant risk factor for SGBV with an odds ratio of 2 and 40% of controls exposed. At a ratio of 1:2 cases to controls and 80% power, we obtained a sample size of 324 (108 cases and 216 controls). Using a line list of SGBV cases from the ten health facilities, we randomly selected 108 SGBV cases for interviews. We visited the cases in the community for interviews with the help of social workers and community health workers. For every case, we identified two neighbourhood-matched control AGYW. The controls were identified from neighbour households of cases. We interviewed one control in each household. If more than one controls was available in a given household, we used a random number to select one for interviews.

We adopted the recent Uganda’s Violence against Children (VAC) Survey (2018) questionnaire, (4) and tailored it to our study. We interviewed cases and controls to obtain information on socio-demographics, risk factors for SGBV, care and services received, economic activities undertaken during the COVID-19 period by the participant and their immediate family place of residence before and during the COVID-19 period, school attendance before and during the COVID-19 period, household size, household members living in the house, household members’ occupations, day and night-time activities of the case, parental supervision, and others. To determine the factors associated with SGBV, we conducted conditional logistic regression analysis.

### Key informant interviews to identify factors associated with SGBV during the pandemic

We interviewed the six GBV focal persons who manage SGBV cases as key informants at the six facilities in which the focal person was present during data collection. Using a predesigned key informant guide that had open ended questions, we asked about possible root causes of increased SGBV cases in the region and explored challenges related to high rates of teenage pregnancies, management of SGBV during the pandemic, and availability of drugs such as emergency contraceptives and PEP.

The interviews were carefully recorded and transcribed, followed by a systematic content analysis approach. The transcriptions were coded to identify recurring themes, which were then organized into broader thematic categories. The results were summarized by extracting relevant information and including verbatim quotes to support the quantitative findings.

### Ethical approval and consent to participate

The Uganda Public Health Fellowship Program under which this evaluation was conducted is part of the National Rapid Response Team, and has been granted permission by the Ministry of Health to access and analyse surveillance data in the District Health Information System-2 and other data such as survey and field investigation data to inform decision making in the control and prevention of outbreaks and public health programming. Additionally, the MOH has also granted the program permission to disseminate the information through scientific publications.

In agreement with the International Guidelines for Ethical Review of Epidemiological Studies by the Council for International Organizations of Medical Sciences (1991) and the Office of the Associate Director for Science, CDC/Uganda, it was determined that this activity was not human subject research and that its primary intent was public health practice or disease control activity (specifically, epidemic or endemic disease control activity). All experimental protocols were approved by the US CDC human subjects review board and the Uganda Ministry of Health and have been performed in accordance with the Declaration of Helsinki. This activity was reviewed by CDC and found consistent with applicable federal law and CDC policy. We obtained a CDC e-clearance (Project ID: 0900f3eb81e95e4d, Accession #: CGH-FETPT-2/17/22-95e4d) “All of the reviews have been completed for your project. You may now work on this project” on 4^th^, Mar, 2022.

Written informed consent was obtained from the participants before the start of each interview including the health care workers and the interviewed AGYW. We obtained written informed consent from the parents and guardians of the AGYW that were below the age of 18 years before interviews and assent from these girls. We also sought permission from the district health officials and heads of the health facilities. During data collection, respondents were assigned unique identifiers instead of names to protect their confidentiality.

## Results

### Description of SGBV cases, Mid-Eastern Region, Uganda, March 2020-December 2021

We identified 389 SGBV cases among AGYW at the ten selected health facilities in Tororo and Busia Districts from March 2020 to December 2021. Their mean age was 16.4 years (SD± 1.61: range, 10-24 years) and most (350; 90%) were 15-19 years of age. Two thirds (214; 67%) were pregnant at the time their SGBV event was recorded in the facility register. Nearly half (180; 46%) presented to the facility >72 hours after the event. Among the 209 who presented within 72 hours, only 8 (4%) received PEP (Table 1).

**Table 1:** Characteristics of sexual gender-based violence cases managed at health facilities during the COVID-19 period in Mid-Eastern Region, 2020-2021.

| Variable | n=389 | (%) |
| --- | --- | --- |
| **Age** |  |  |
| 10-14 | 25 | (6.4) |
| 15-19 | 350 | (90) |
| 20-24 | 14 | (3.6) |
| **Marital status** |  |  |
| Married | 198 | (51) |
| Not married | 191 | (49) |
| **HIV test done** |  |  |
| Yes | 321 | (83) |
| No | 65 | (17) |
| **Followed up by HCW after initial visit at least once** |  |  |
| Yes | 16 | (4.1) |
| No | 373 | (96) |
| **Followed up by HCW after initial visit at least 4 times (as recommended)** |  |  |
| Yes | 0 | (0) |
| No | 389 | (100) |
| **Pregnant** |  |  |
| Yes | 214 | (67) |
| No | 107 | (33) |
| **Time between event and presentation for care** |  |  |
| <24 hours | 97 | (25) |
| 24-72 hours | 112 | (29) |
| >72 hours | 180 | (46) |
| **PEP among person who presented <72 hrs (n=209)** |  |  |
| Yes | 8 | (3.8) |
| No | 201 | (96) |

\*HCW- Health care worker

### Description of sexual gender-based violence cases considered for the case-control study

Of the 108 cases interviewed, 79 (73%) reported being physically forced into sex while 29 (27%) reported that they were pressured into sex through harassment and threats. Most (88; 82%) reported that the episode was their first SGBV experience during COVID-19 period. Among 73 (68%) who agreed to share information about the perpetrator, 29 (40%) reported it was a friend and 17 (23%) said it was the neighbour. The majority (71; 67%) reported to the health facility after >72 hours; of these, 42 (58%) said that they delayed reporting due to feelings of social stigma about their SGBV experience. The most reported adverse outcome from the episode was unwanted pregnancy (38; 35%) (Table 2).

**Table 2:** Characteristics of sexual gender-based violence cases included in the case control study during the COVID-19 period in Mid-Eastern Region, 2020-2021.

| Variable | N=108 | (%) |
| --- | --- | --- |
| **Type of SGBV** |  |  |
| Physically forced sex | 79 | (73) |
| Pressured into sex through harassment and threats | 29 | (27) |
| **First time experience of SGBV during COVID-19** | 88 | (82) |
| **Time of Sexual event** |  |  |
| Day | 23 | (21) |
| Evening | 21 | (19) |
| Night | 44 | (40) |
| Declined to answer | 20 | (19) |
| **Type of perpetrator (among 73 providing information)** |  |  |
| Friend | 29 | (40) |
| Neighbor | 17 | (23) |
| Relative | 9 | (12) |
| Others | 18 | (25) |
| **Where the incident occurred** |  |  |
| On the way somewhere | 25 | (21) |
| Friends place | 25 | (23) |
| Home | 26 | (24) |
| Relative home | 11 | (10) |
| Workplace | 2 | (1.9) |
| Not comfortable to mention the place | 20 | (19) |
| **Adverse outcomes** |  |  |
| Unwanted pregnancy | 38 | (35) |
| Depression | 20 | (19) |
| Anxiety | 12 | (11) |
| STI | 7 | (6.5) |
| Others | 14 | (13) |
| **Reported they received emergency contraception** | 35 | (32) |
| **Time from SGBV event to presentation to health facility** |  |  |
| Immediately | 18 | (27) |
| 1-3 days | 18 | (27) |
| >3 days | 72 | (67) |
| **Self-reported they received PEP from health facility or other site (among 102 who provided a response)** | 35 | (34) |
| **Self-reported they received PEP from health facility or other site (among 36 who presented <3 days after event)** | 20 | (56) |
| **Reasons for reporting to the health facility > 3 days (n=72)** |  |  |
| Social stigma | 42 | (58) |
| Fear of retaliation | 13 | (18) |
| Lack of awareness | 13 | (18) |
| Others | 4 | (5.6) |

\*PEP; Post Exposure Prophylaxis, STI: Sexually Transmitted Infection

### Factors associated with sexual gender-based violence among adolescent girls and young women during COVID-19, Tororo and Busia districts, Mid-Eastern Uganda

In bivariate analysis, having older siblings [cOR=1.7, 95% CI: 1.04-2.8], ever being pregnant before the COVID-19 period [cOR=2.5, 95% CI: 1.6-4.0], ever being sexually violated before the COVID-19 period [cOR=8.1, 95% CI: 4.1-16], and ever being involved in sex work [cOR=3.4, 95% CI: 2.1-5.6] were associated with SGBV. In contrast, having a family that was able to provide all basic needs was protective [cOR=0.33, 95% CI: 0.20-0.56] (Table 3). Age, being in school before COVID-19 period, level of education, ever having had children, knowledge and use of family planning, and family change in economic status during the COVID-19 period were not significantly associated with SGBV.

**Table 3:**
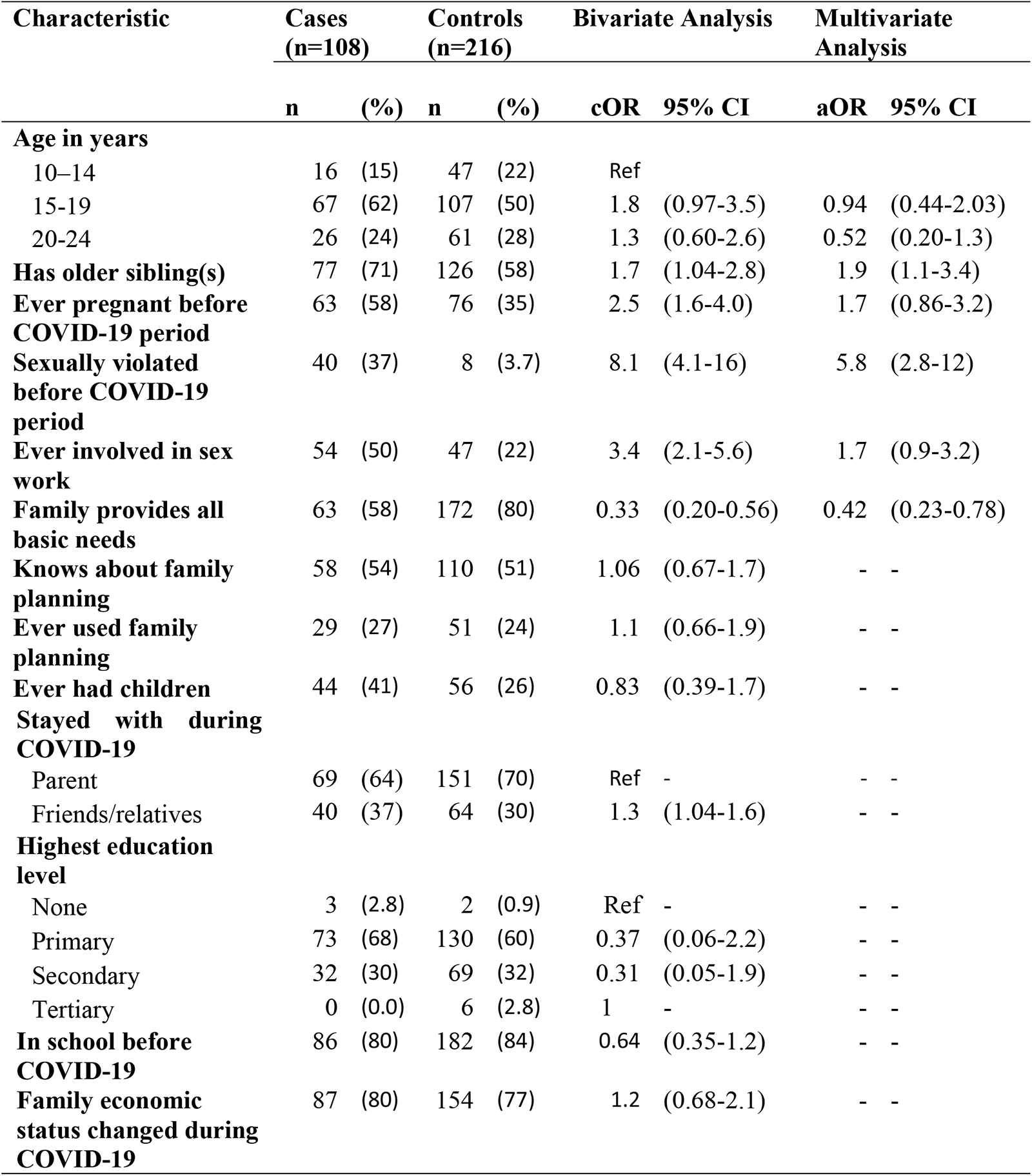
Characteristics of cases and controls and their association with Sexual Gender Based Violence during the COVID-19 period in Mid-Eastern Region, 2020-2021.

| Characteristic | Cases<br>(n=108) |  | Controls<br>(n=216) |  | Bivariate Analysis |  | Multivariate Analysis |  |
| --- | --- | --- | --- | --- | --- | --- | --- | --- |
|  | n | (%) | n | (%) | cOR | 95% CI | aOR | 95% CI |
| **Age in years** |  |  |  |  |  |  |  |  |
| 10–14 | 16 | (15) | 47 | (22) | Ref |  |  |  |
| 15-19 | 67 | (62) | 107 | (50) | 1.8 | (0.97-3.5) | 0.94 | (0.44-2.03) |
| 20-24 | 26 | (24) | 61 | (28) | 1.3 | (0.60-2.6) | 0.52 | (0.20-1.3) |
| **Has older sibling(s)** | 77 | (71) | 126 | (58) | 1.7 | (1.04-2.8) | 1.9 | (1.1-3.4) |
| **Ever pregnant before COVID-19 period** | 63 | (58) | 76 | (35) | 2.5 | (1.6-4.0) | 1.7 | (0.86-3.2) |
| **Sexually violated before COVID-19 period** | 40 | (37) | 8 | (3.7) | 8.1 | (4.1-16) | 5.8 | (2.8-12) |
| **Ever involved in sex work** | 54 | (50) | 47 | (22) | 3.4 | (2.1-5.6) | 1.7 | (0.9-3.2) |
| **Family provides all basic needs** | 63 | (58) | 172 | (80) | 0.33 | (0.20-0.56) | 0.42 | (0.23-0.78) |
| **Knows about family planning** | 58 | (54) | 110 | (51) | 1.06 | (0.67-1.7) | - | - |
| **Ever used family planning** | 29 | (27) | 51 | (24) | 1.1 | (0.66-1.9) | - | - |
| **Ever had children** | 44 | (41) | 56 | (26) | 0.83 | (0.39-1.7) | - | - |
| **Stayed with during COVID-19** |  |  |  |  |  |  |  |  |
| Parent | 69 | (64) | 151 | (70) | Ref | - | - | - |
| Friends/relatives | 40 | (37) | 64 | (30) | 1.3 | (1.04-1.6) | - | - |
| **Highest education level** |  |  |  |  |  |  |  |  |
| None | 3 | (2.8) | 2 | (0.9) | Ref | - | - | - |
| Primary | 73 | (68) | 130 | (60) | 0.37 | (0.06-2.2) | - | - |
| Secondary | 32 | (30) | 69 | (32) | 0.31 | (0.05-1.9) | - | - |
| Tertiary | 0 | (0.0) | 6 | (2.8) | 1 | - | - | - |
| **In school before COVID-19** | 86 | (80) | 182 | (84) | 0.64 | (0.35-1.2) | - | - |
| **Family economic status changed during COVID-19** | 87 | (80) | 154 | (77) | 1.2 | (0.68-2.1) | - | - |

In multivariable analysis, experiencing sexual violence before COVID-19 [aOR=5.8, 95% CI: 2.8-12] and having older siblings [aOR=1.9, 95% CI: 1.1-3.4] remained significantly associated with SGBV during COVID-19 period. The family’s ability to provide all basic needs [aOR=0.42, 95%: CI 0.23-0.78] was protective. While the odds of SGBV decreased with increasing age, the association was not significant (Table 3).

### Qualitative results

Results from the key informant interviews supported these findings.

Most GBV focal persons reported that AGYW who experienced SGBV often presented to the health facility beyond the recommended 72 hours. In many cases, these survivors were only identified during their antenatal care visits, often due to pregnancies resulting from the SGBV incidents. Consequently, they were unable to receive PEP and emergency oral contraceptives, despite the availability of such medicines. The management of SGBV cases posed a significant challenge during the COVID-19 period, as expressed by one key informant*: “The management of the SGBV cases was a big challenge during the COVID-19 period. Most girls presented here very late, some already pregnant, and whenever you asked them why they never came early, they reported they feared the perpetrators. You had nothing to do for them apart from counseling” (Key Informant II)*.

The key informant respondents’ highlighted significant parenting issues during the COVID-19 period, noting that many young people had unrestricted movement due to school closures and were frequently seen walking at night. This lack of parental supervision and control was a concern, as it potentially exposed girls to high-risk situations that could contribute to sexual gender-based violence. One key informant stated, “*I think parents did not play a good role during the COVID-19 lockdown. You know children were very idle and had nothing to do*. *You could find a group of girls and boys walking and laughing in the evenings as if they had no chores at home. Also, in this community, it is common to find young people moving at night, and this is even worse when there is a party” (Key Informant IV)*.

Five out of six GBV focal persons highlighted that the families faced challenges in meeting their basic needs. The COVID-19 pandemic led to widespread poverty due to disrupted businesses. Consequently, many parents were unable to provide adequate support to their families. The resulting lack of necessities such as food may have compelled young people including the AGYW to seek necessities from other sources. *Key informant IV stated, “During COVID-19, there was a lot of poverty due to disrupted businesses and closure of schools. As a result, most parents were unable to support their children. Such could have forced girls to look for necessities such as food and clothing from elsewhere”*

## Discussion

During the COVID-19 period, sexual gender-based violence (SGBV) became a pervasive and often overlooked challenge, perpetuating a disturbing silence that hindered effective intervention and support to the survivors. Many reported cases were pregnant at the time their SGBV experience was recognized and recorded. Shockingly, close to half of the survivors delayed reporting their experiences, making them ineligible for timely access to Post Exposure Prophylaxis (PEP). Even among those who reported earlier, the utilization of PEP services remained alarmingly low. Previous experiences of sexual violence and having older siblings were found to increase the likelihood of SGBV, while having sufficient family resources was a protective factor. Key informant interviews highlighted delays in seeking post-violence services, parenting challenges, and the lack of basic necessities as contributing factors to SGBV among AGYW during the COVID-19 period. These findings underscore the pressing need for comprehensive interventions to address and prevent SGBV, particularly in times of crisis.

Our findings indicate that less than half (34%) of SGBV victims self-reported to have received PEP. The low uptake of PEP among the exposed persons raises concerns about their risk of HIV acquisition. PEP is a critical intervention that can significantly reduce the risk of HIV transmission after a potential exposure, such as sexual assault (11). These findings were in line with a study done in Uganda where analysis of health information system (HMIS) data revealed 18% reduction on PEP uptake despite the increased sexual violence reports during COVID-19 period (6). PEP is available in public and private health care facilities in Uganda and must be given within 72 hours of the event to be effective (12). Low PEP uptake raises significant concerns about the provision of appropriate healthcare services to survivors of SGBV during COVID-19 pandemic. We identified many missed opportunities for preventing HIV infection among AGYW who experienced SGBV in Mid-Eastern Region. Several pandemic-related factors such as movement restrictions, limitations in resources and personnel, shifting priorities, and changes in service delivery models (13) may have impacted the availability and accessibility of healthcare services, including PEP (14).

Even among the AGYW who presented early, few received PEP. However, even among those who received PEP, it was often inappropriately given, highlighting a knowledge gap on PEP use among the health care workers and the users. Due to data gaps in the SGBV health facility registers, we could not verify the appropriateness of PEP use for all reported incidents. Similar findings were noted in a study done in Uganda to compare post-rape care before and during COVID-19 using national Health Management Information System (HMIS) and Uganda Child Help Line (UCHL) data. In this study, 50% of cases received PEP beyond the recommended 72 hours (15). This highlights the need for flexible methods of service delivery to increase timely access and utilisation of PEP among SGBV survivors. These might include community based approaches such as “off-facility PEP medication delivery” and “PEP hotlines” (16). These community-based approaches were piloted in rural populations in Kenya and Uganda; results showed increased uptake and completion of PEP in the population (16). Emphasizing PEP guidelines on use within 72 hours among health care workers may be necessary to facilitate more appropriate use since PEP is effectiveness is time bound.

Interviews with the GBV focal persons, as well as the high rate of pregnancy among AGYW identified as SGBV survivors from the clinic registers, suggested that many AGYW presented because of their pregnancy rather than the SGBV event itself. AGYW cited fears of the perpetrator and stigma as reasons they did not present immediately. Given that many SGBV events will not result in pregnancy, the high frequency of pregnancy among SGBV survivors who do report suggests that the reports described in this paper are the tip of the iceberg. Many women who do not become pregnant because of the SGBV likely simply do not report. Fear of public stigma is a well-documented factor that hinders patients from accessing health care services (9,10). Unfortunately, social stigma related to SGBV is known to worsen physical and psychological health impacts (18) and may increase the risk of additional violence (19). Efforts are needed to support AGYW to report SGBV and enable adequate and timely care to prevent HIV and unwanted pregnancies. The UCHL 116, which allows reporting by phone, was found to be a more acceptable channel to report sexual violence than reporting at health facilities (20). UCHL was implemented by Ministry of Gender Labour and Social Development (MGLSD) to improve reporting of child abuse and sexual violence. However, there is still low awareness of the UCHL, and a need for more advocacy (20). Active case-finding for SGBV survivors may be necessary to ensure appropriate care and support to the affected persons.

Having experienced SGBV before the COVID-19 period was associated with SGBV during COVID-19. Repeated sexual violence during COVID-19 suggests that affected girls may be continually exposed to the same perpetrator(s). Other studies have also found that individuals who are sexually victimised were likely to be re-victimised (15, 16,17). Both psychosocial support and ways to protect the once identified survivors such as establishing peer support groups to create safe spaces for victims (20), economic empowerment initiatives, community mobilization (22) may be necessary to prevent repeated SGBV experiences. In addition, there is need to strengthen the parents’/guardians’ daughter relationship to enable the girl’s open up to parents in case of SGBV through evidence based interventions such as community parenting programs and whole family support programming (23) (24).

The association between having older siblings and an increased risk of sexual gender-based violence (SGBV) observed in our study was not clear. However, it may simply reflect a younger age of the SGBV victim compared to non-SGBV survivors; age was found to be negatively associated with SGBV in our study, with the odds of association decreasing as age increased. Other factors could be related to power dynamics (25) or older siblings having experienced or witnessed SGBV, perpetuating a cycle of violence within the family (26). Exposure to violence can impact their attitudes towards relationships and sexual behavior, potentially leading to higher incidences of SGBV among younger siblings (27).Interventions that promote healthy sibling relationships and fostering positive role modeling may be necessary to address SGBV. Nevertheless, further research is needed to gain a deeper understanding of the underlying mechanisms involved and provide a potential explanation for this association.

We found that having a family providing all the basic needs to AGYW was protective against SGBV during COVID-19. Unfortunately, as a result of COVID-19 measures like the closure of bars, shops, saloons, and markets and movement restrictions (28), many families were unable to provide basic needs for their children (21, 22). Efforts to support vulnerable AGYW with basic needs like food, clothing, and shelter among others may be necessary during future pandemics or similar situations to prevent SGBV.

## Limitations

Our study was subject to several limitations. Health facility data were incomplete, limiting what we could analyse. We found irregularities with documentation in health facility registers, especially information on dates of incidence and follow-up services, and data on repeated episodes of SGBV. We could not tell from the data if the persons who were offered PEP accepted or completed the dosage. In addition, information on the first point of care was missing, and thus we could not establish if the cases who were recorded as pregnant were identified only during their antenatal clinic visits. We used the health facility data to identify the cases, and thus our data certainly represent an underestimate of the problem as well as reflecting possible bias if the survivors who did not present to the facility differ from those who do. Also, given the sensitivity of the subject and self-reporting of the data, some cases did not respond to questions related to the perpetrator resulting into potential bias.

## Conclusion

Sexual gender-based violence negatively affected the lives of many adolescent girls and women during the COVID-19 pandemic. Most survivors did not present in time for HIV exposure prophylaxis and therefore efforts to improve access to services through community education and engagement with AGYW is key for PEP uptake when needed. Public health programs in the future may need to focus on identifying or supporting known survivors of SGBV and those who are socioeconomically vulnerable and identifying approaches to protect them, especially during school closures or other events that leave them close to home.

## Availability of data and materials

The datasets upon which our findings are based belong to the Uganda Public Health Fellowship Program, Ministry of Health, Uganda. For confidentiality reasons, the datasets are not publicly available. However, the data sets can be made available upon reasonable request from the corresponding author and with permission from the Uganda Public Health Fellowship Program.

## List of abbreviations

SGBV: Sexual Gender-Based Violence
GBV: Gender-Based Violence
AGYW: Adolescent Girls and Young Women
UPHIA: Uganda Population Impact Assessment
DHIS2: District Health Information System
PEP: Post Exposure Prophylaxis
VAC: Violence Against Children
MoH: Ministry of Health
COVID-19: Corona Virus Disease

## Disclaimer

The findings and conclusions in this report are those of the author(s) and do not necessarily represent the official position of the U.S. Centers for Disease Control and Prevention.

## Authors’ contributions

P.M took the lead in conceptualizing the study idea, data analysis, writing and editing of the manuscript. P.M, J.C, contributed to the development of the proposal and data collection tools, and P.M, B.K, RM contributed to epidemiological data collection and formal analysis of data, P.M, R.M, B.K, D.K, L.B, A.R.A contributed to interpretation of findings investigation, writing, editing, and reviewing of the manuscript. All authors contributed to the write-up and read and approved the final manuscript.

## Acknowledgments

We appreciate the health facilities and the data collection teams, especially the health facility GBV focal persons, for the support rendered to interview the cases. We commend the AGYW who participated and shared their experiences to inform the factors associated with SGBV during the COVID-19 period. I acknowledge Julie R. Harris and Rose Apondi for their technical support rendered to me during the inception of the idea and review of the manuscript.

